# Intentions to get vaccinated against Monkeypox in Healthcare workers in France and Belgium correlates with attitudes toward COVID-19 vaccination

**DOI:** 10.1101/2022.08.25.22279205

**Authors:** Amandine Gagneux-Brunon, Nicolas Dauby, Odile Launay, Elisabeth Botelho-Nevers

**Affiliations:** Centre International de Recherche en Infectiologie, Team GIMAP, Univ Lyon, Université Jean Monnet, Université Claude Bernard Lyon 1, Inserm, U1111, CNRS, UMR530; CIC INSERM 1408 Vaccinologie, CHU de Saint-Etienne; Chaire PREVACCI, Université Jean Monnet, Saint-Etienne, France; Department of Infectious Diseases, CHU Saint-Pierre - Université Libre de Bruxelles (ULB), Brussels, Belgium; School of Public Health, université libre de Bruxelles (ULB); Université de Paris, Inserm CIC 1417, Assistance publique – Hôpitaux de Paris, hôpital Cochin, 75679 Paris, France France

**Keywords:** Monkeypox, vaccination, vaccine hesitancy, healthcare workers, attitudes

## Abstract

**Background:** Front-line healthcare workers (HCWs) could be at-risk for Monkeypox infections. Vaccine hesitancy also affects HCWs and has an impact on their own attitudes toward vaccination. In the context of the exhaustion due to COVID-19 pandemic, we aimed to evaluate intentions to get vaccinated against Monkeypox in HCWs in France and Belgium.

**Methods:** We performed a cross-sectional study (snowball sampling) using a self-administered online questionnaire to evaluate intentions to get vaccinated against Monkeypox in HCWs if a recommendation for HCWs vaccination was made. We compared demographics characteristics, vaccine readiness, eagerness for COVID-19 vaccine, and confidence in HCW with Chi-square tests, student-t and performed a binary regression.

**Results:** Amon the 397 respondents, if a specific recommendation was made for HCWs vaccination against Monkeypox was made, 55.4 % will probably get the vaccine, while 79 % would accept the vaccine if recommended to the general population. COVID-19 vaccine eagerness and having concerns about Monkeypox epidemics were associated with favorable attitude toward Monkeypox vaccination in HCWs with respective adjusted odds ratio and 95 % Confidence Interval 2.5 (1.03-6.1), 2.6 (1.3-5.3). Forty-four HCWs (11 %) self-identified as at-risk for Monkeypox infections.

**Conclusion:** Acceptance of Monkeypox vaccination in HCWs is probably moderate, HCWs are probably complacent and did not perceive the risk of Monkeypox infections in the context of professional exposure.

---

Europe is currently the epicenter of the Monkeypox epidemics. On the 09^th^ of August 2022, 17,897 cases of Monkeypox have been identified [1]. Forty-eight infections occurred in health care workers (HCWs) with no identified occupational exposure. In Israel, a case was reported in a physician after occupational exposure to a patient with Monkeypox in spite of the use of protective equipment [2]. World Health Organization (WHO) recommends immunization for HCWs at-risk for occupational exposure [3]. However, in the context of limited vaccines supply, national authorities should consider whether HCWs may be at risk of repeated exposure and the possible nature of the exposure. In the United Kingdom, face to an increased number of cases, pre-exposure prophylaxis is recommended to staff working in high consequence infectious diseases units, Monkeypox patients-dedicated units and in outpatients sexual health clinics [4].

Vaccine hesitancy affects HCWs and have an impact on their personal vaccination status [5]. After two years of COVID-19 pandemic and vaccination, attitudes toward vaccination against a new emerging infection is not determined, particularly in France where mandates were required to achieve high vaccine coverage in HCWs. We carried out an anonymous on-line survey from the 15^th^ of June 2022 to the 8^th^ of August 2022, by snowball sampling in France and Belgium, to evaluate attitudes toward Monkeypox vaccination. Among the 690 responders, 397 were HCWs (mean age 43.3 ± 12 years, 260/397 women). We aimed to evaluate intentions to get vaccinated in HCWs and to identify factors associated with this attitude (demographic characteristics, 7C vaccination readiness [6], confidence in government, public health agencies, pharmaceutical companies, medias, and colleagues, and COVID-19 vaccine eagerness).

## Intentions to get vaccinated against Monkeypox in Healthcare workers

If a specific recommendation was done for HCWs, 99 (30.5 %) of the 397 respondents would get the vaccine as soon as possible, 121 (24.9 %) would probably get vaccinated, 88 (22.2 %) were undecided, 49 (12.3) would probably not get the vaccine, and 40 (10.1%) would certainly not get the vaccine. Overall, two hundred and twenty (55.4 %) would accept vaccination. Among the 328 who reported their vaccine status against smallpox, 127 (38.7 %) declared that they received smallpox vaccine, smallpox vaccine history had no impact on intentions to receive the vaccine. Among the 177 HCWs who did not intend to get vaccinated if vaccination was specifically recommended to HCWs, 96 (54.2%) will accept to get the vaccine if the epidemics spread to the general population. In this context, 314 (79.1 %) of the HCWs responders would accept the vaccine. If vaccination against Monkeypox would became recommended in the general population, 314 responders (79 %) would accept to get vaccinated.

## Factors associated with intention to get vaccinated against Monkeypox

Forty-four (11 %) responders felt at-risk of Monkeypox infection, and 87 (21.9 %) expressed concerns about the current Monkeypox epidemics. In the context of a specific recommendation for HCWs, the proportion of HCWs with intention to get vaccinated was significantly greater in physicians and pharmacists (65.3 % versus 46.6 % nurses and assistant Nurses or 43 % in other professions-midwives, physiotherapists, etc…-). The differences between physicians/pharmacists and nurses and assistant nurses in attitudes toward Monkeypox vaccination were also observed in the eventuality of an extension of the vaccination to the general population: 84.7 % in physicians and pharmacists, 70.7 % in nurses and assistant nurses, 77.2 % in other HCWs. In univariate analysis, intentions to get vaccinated against Monkeypox if the vaccine was specifically recommended for HCWs were associated with attitudes toward vaccines in general assessed by the 7C vaccination readiness, and eagerness for COVID-19 vaccination (Table 1). HCWs who reported multiple sexual partners were not more prone to get a vaccine against Monkeypox. After adjustment on age, 7C vaccination readiness, intention to get vaccinated against Monkeypox was associated with COVID-19 vaccine eagerness (defined by vaccination before eligibility or as soon as HCWs became eligible). In addition, confidence in government, public health agencies, medias, and colleagues (assessed by a 30-point score) was associated with intentions to get vaccinated against Monkeypox, if vaccination was specifically recommended in HCWs. In this multivariable regression, nurses and assistant nurses were not less prone to intend for vaccination against Monkeypox than physicians and pharmacists. Concerns about Monkeypox epidemics and COVID-19 vaccine eagerness were also main predictors of intentions to get vaccinated against Monkeypox if the vaccination recommendation was extended to the general population.

**Table 1:**
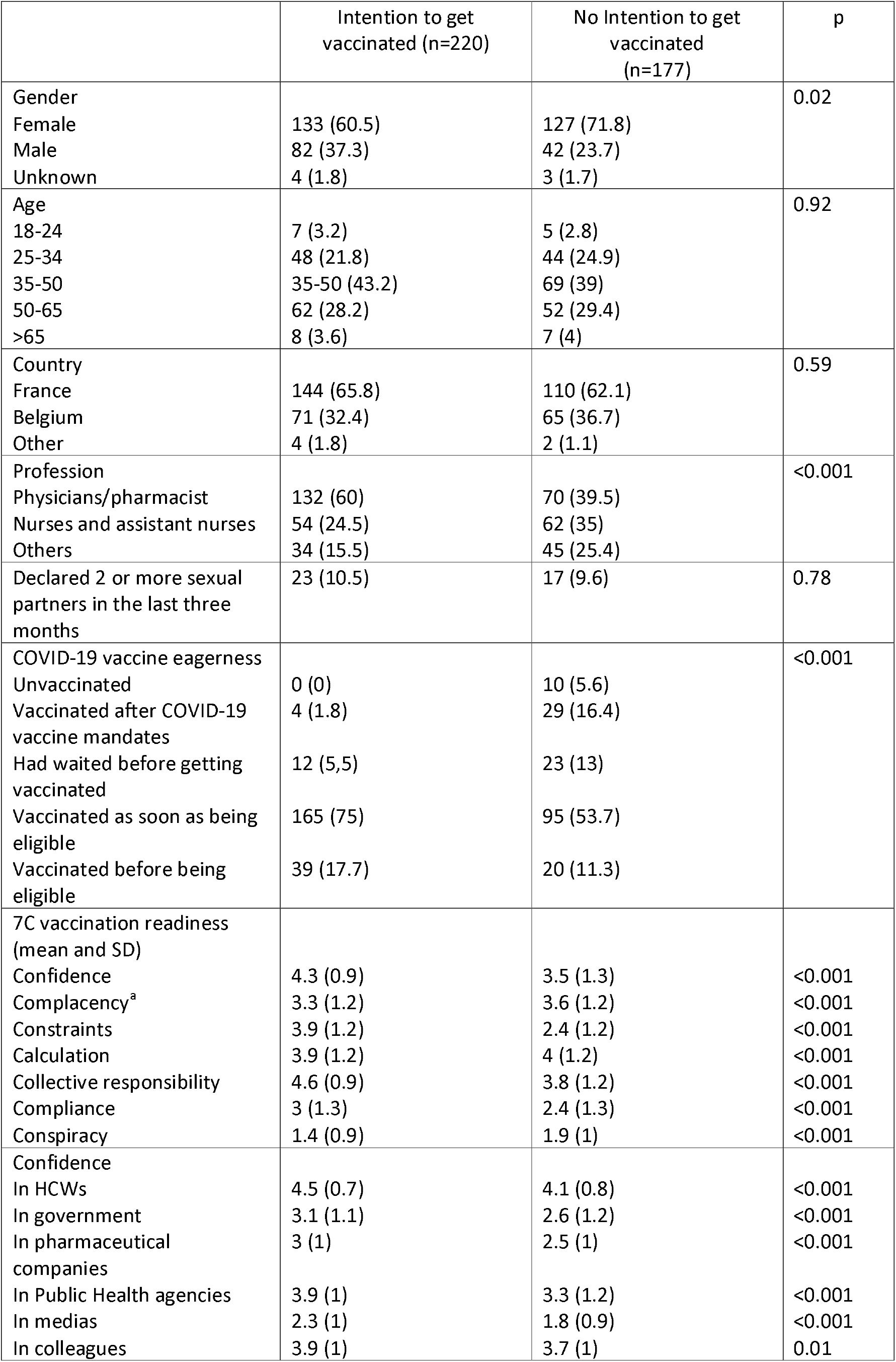
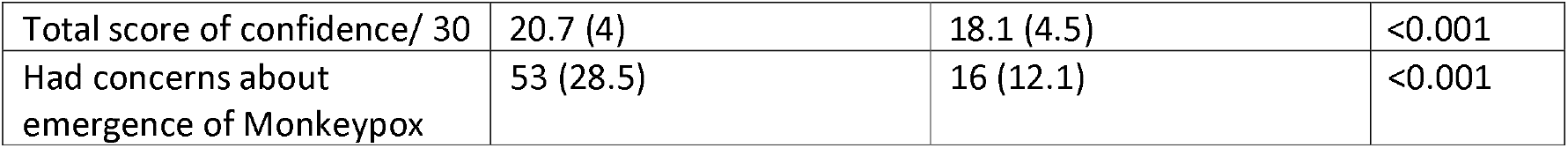
Characteristics of the 397 respondents divided in two groups: HCWs with intention to get vaccinated, HCWs without intention to get vaccinated against Monkeypox. number and (%) for categorical variables, and mean and (standard deviation) for continuous variables, Univariate analysis was performed with chi square tests for categorical variables and student t-test for continuous variables. ^a^ Higher is the score, less the responders is complacent.

**Table 2:**
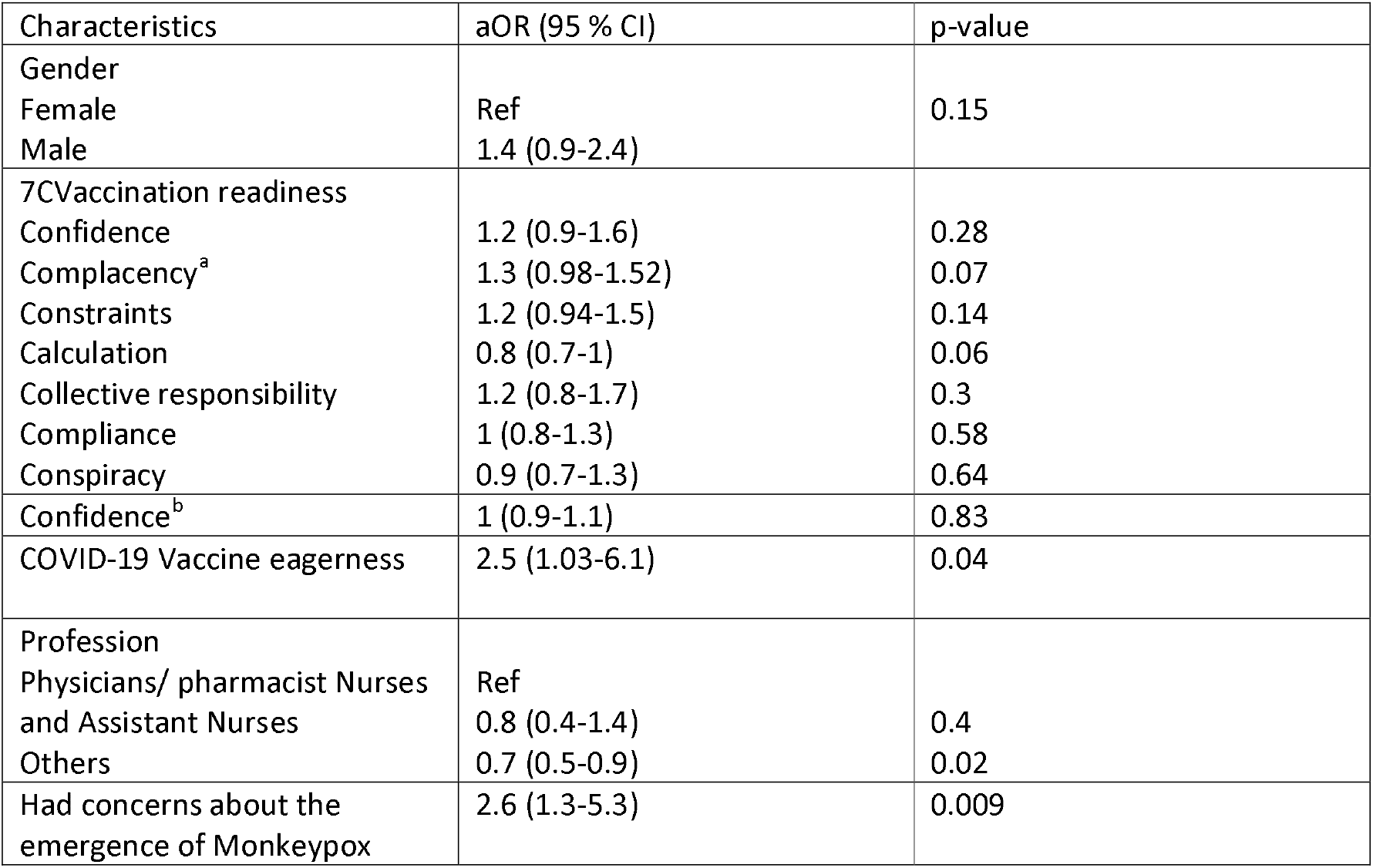
Factors associated with intention to get vaccinated against Monkeypox in healthcare workers. obtained with multivariable binary regression model adjusted gender, profession, COVID-19 vaccine eargerness; 7C-vaccination readiness, confidence, and concerns about Monkeypox epidemics. aOR adjusted odds ratio ^a^Higher is the score, less the responders is complacent, ^b^a confidence 30-point score was obtained adding the 6 items 5 likert scale results, COVID-19 eagerness was transformed in a binary variable: individuals vaccinated as soon as they were eligible or before being eligible were defined as eager.

Acceptance of the Monkeypox vaccination in HCWs is limited in the case of a specific recommendation for professional exposure, but is not very far from the acceptance rate observed in men having sex with men (MSM) in the Netherlands if the vaccine become recommended in the general population [7]. Different hypotheses can be made. First, HCWs could suffer from pandemic fatiguespecifically about vaccine recommendations, notably in France, where COVID-19 vaccine mandates were implemented. Secondly, HCWs could be complacent about Monkeypox infections, as a minority of them felt at-risk for Monkeypox infection. At the beginning of the COVID-19 pandemic, self-perceived risk for infection was an important predictor of the attitudes toward a vaccine [8]. A communication focused on mild infections mainly affecting MSM generate feelings of complacency in HCWs [9]. Complacency for Monkeypox is probably a great barrier for vaccine acceptance. In MSM, in the Netherlands, perceived risk of being infected and risky sexual behaviors did not predict attitudes toward vaccination, while having concern about being infected was associated with intention to get vaccinated [7]. Then, in the context of the epidemics, the large use of personal protective equipment may also give to HCWs a sense of security. However, case of transmission probably through contact with contaminated bedding was previously reported suggesting that front-line HCWs could be at-risk of Monkeypox infection [10]. Our observation may suggest that communicating on potential professional exposure to Monkeypox in HCWs is required. The main limitations of our work are the lack of representativeness of our sample of the HCWs in France and Belgium and the relatively small sample size. Physicians and pharmacists were overrepresented, and due to more favorable attitudes toward vaccines in physicians and pharmacists in general, intentions of getting vaccinated against Monkeypox in HCWs were probably overestimated. Lower intentions to get vaccinated in nurses and assistant nurses than in physicians and pharmacists in univariate analysis but not in multivariate analysis are probably due to less confidence, more complacency, lower level of knowledge and more negative attitudes toward vaccines [5]. Highly prevalent vaccine hesitancy in nurses and assistant nurses is particularly challenging in the context of infectious emerging diseases as their close contacts with infected patients are frequent [11]. In addition, it is probable that an evolution in attitudes toward Monkeypox vaccination will be observed in parallel to the evolution of the epidemics. In the regression model, the 7Cs components were not independent predictors of attitudes toward Monkeypox vaccination. This observation may suggest that a specific model could be developed to predict attitudes toward vaccines and measure vaccine hesitancy in HCWs particularly in the context of professional immunizations [5]. In conclusion, this first survey about acceptability of Monkeypox vaccine among HCWs showed a modest acceptability rate if specific recommendations were implemented and highlights the need to address the low perceived risk of occupational transmission.

## Data Availability

All data produced in the present study are available upon reasonable request to the authors

